# Parkinson’s Disease Detection using XGBoost and Machine Learning

**DOI:** 10.1101/2023.10.23.23297369

**Authors:** Dheiver Francisco Santos

## Abstract

This article explores the application of machine learning, specifically the XGBoost algorithm, for the early detection of Parkinson’s disease. Parkinson’s disease is a prevalent neurodegenerative condition that poses diagnostic challenges, particularly in its early stages. To address these challenges, a comprehensive dataset, including vocal frequency measurements, audio analyses, and demographic data, is employed. Data preprocessing techniques, including Min-Max scaling and Synthetic Minority Over-sampling Technique (SMOTE), are applied to prepare the dataset. The XGBoost model is then developed and fine-tuned to achieve an accuracy of approximately 93.33% in detecting Parkinson’s disease. The model exhibits high precision, recall, and F1-Score, making it a valuable tool for early disease detection in the healthcare domain. The study highlights the transformative potential of machine learning in improving patient outcomes and healthcare efficiency.

## Introduction

Parkinson’s disease is a pervasive and debilitating neurodegenerative condition, affecting millions of individuals globally. This complex disorder manifests through a myriad of symptoms, including tremors, muscle rigidity, and difficulties with movement, profoundly affecting the quality of life of those it afflicts (Gomathy et al., 2023; Kulkarni et al., 2023). Timely and precise detection of Parkinson’s disease holds the key to enabling effective interventions and treatments, thereby mitigating the progression of the disease. Machine learning, with its exceptional capacity to analyze large and intricate datasets, presents a promising avenue for early disease detection. In this article, we delve into the application of machine learning, particularly the XGBoost algorithm, for the detection of Parkinson’s disease, leveraging clinical data and biomedical information (Hossain et al., 2023). Our aim is to create a model that can accurately identify the presence of Parkinson’s disease based on specific features.

Parkinson’s disease has been a challenging condition to diagnose accurately, especially in its early stages when symptoms may be subtle and easily overlooked (Zhang et al., 2023). The conventional diagnostic methods have their limitations, underscoring the need for innovative and data-driven approaches. A comprehensive dataset tailored to the task of Parkinson’s disease detection forms the foundation of our study, capturing a wealth of patient-specific information. This dataset encompasses critical data such as vocal frequency measurements, audio analyses, and demographic details (Pinto et al., 2023). The binary “status” feature within the dataset is instrumental in categorizing individuals as either diagnosed with Parkinson’s disease or not, offering a clear distinction for model training and evaluation.

To ensure that our dataset was well-prepared for the task at hand, our methodology involved meticulous data preprocessing steps (Gomathy et al., 2023). Feature normalization through Min-Max scaling was employed, aligning the dataset’s diverse features within a consistent range. This standardization is essential for facilitating model convergence and optimizing its performance. Additionally, our dataset was thoughtfully partitioned into distinct training and testing sets (Kolekar et al., 2023). The training set served as the foundation for the model’s learning, while the testing set facilitated an unbiased evaluation of the model’s performance. Acknowledging the potential for class imbalance within our dataset, we implemented the Synthetic Minority Over-sampling Technique (SMOTE) to rectify this issue. SMOTE significantly improved class balance by oversampling the minority class, reducing the potential for bias in our model’s predictions.

In the wake of this study’s comprehensive methodology, we endeavor to present a robust framework for the early detection of Parkinson’s disease. By harnessing the power of the XGBoost algorithm, a distinguished gradient boosting technique, we aim to demonstrate the potential of machine learning in healthcare. Our model’s accuracy, precision, recall, and evaluation metrics will shed light on its capability to accurately detect Parkinson’s disease (Gomathy et al., 2023). Ultimately, our goal is to contribute to the enhancement of patient outcomes and healthcare efficiency by providing a valuable tool for early disease detection in the realm of Parkinson’s disease.

## Methodology

In this study, we commenced by assembling a comprehensive dataset tailored for Parkinson’s disease (PD) detection. The dataset was thoughtfully designed to encompass an array of patient-specific data, including vocal frequency measurements, audio analyses, and essential demographic information. A pivotal attribute of the dataset is the binary “status” feature, which categorizes individuals into two groups: those diagnosed with PD (assigned a value of 1) and those without (assigned a value of 0). Our data preparation process included thorough data cleansing, the handling of missing values, and ensuring data consistency.

To ensure that the dataset was optimally configured for both model training and evaluation, we undertook a series of data preprocessing steps. Firstly, we employed the Min-Max scaling technique to normalize the features within the dataset, thereby bringing all variables into a uniform range. This normalization process is critical to ensure that the model converges effectively. Secondly, the dataset was thoughtfully partitioned into separate training and testing sets. The training set was employed to facilitate model training, while the testing set served as the foundation for an unbiased assessment of the model’s performance. Additionally, recognizing the potential for class imbalance within our dataset, we introduced the Synthetic Minority Over-sampling Technique (SMOTE) to address this issue. By oversampling the minority class, SMOTE substantially improved class balance, reducing the possibility of bias in the model’s predictions.

The core of our research centered on the development and evaluation of our Parkinson’s disease detection model. We harnessed the capabilities of the XGBoost algorithm, a distinguished gradient boosting technique well-regarded for its classification prowess. With the training data, augmented through SMOTE, our model underwent meticulous training. An iterative process of hyperparameter tuning allowed us to fine-tune key parameters such as learning rate, tree depth, and the number of boosting rounds. This optimization ensured that our model was configured for peak performance. Finally, the trained model was subjected to rigorous evaluation, encompassing various metrics, including accuracy, precision, recall, F1-Score, and the generation of receiver operating characteristic (ROC) and precision-recall (PR) curves. These metrics collectively provided a comprehensive evaluation of the model’s ability to accurately detect Parkinson’s disease.

In addition to developing and evaluating our Parkinson’s disease detection model, we also aimed to enhance its interpretability. Model interpretation is crucial in the healthcare domain, where understanding the factors contributing to predictions is vital. To achieve this, we employed techniques such as feature importance analysis. This analysis enabled us to identify the most influential features in making PD predictions, providing valuable insights into the disease’s diagnostic factors. By delving into feature importance, we contributed to the transparency and interpretability of our model.

Research in healthcare and medical diagnosis necessitates ethical considerations. Throughout our study, we adhered to ethical guidelines and ensured that patient data, whether real or simulated, was handled with utmost care and privacy. We obtained any necessary approvals and consents, and all data handling was in compliance with relevant data protection regulations. This ethical dimension is crucial in maintaining the trust and integrity of our research. It’s important to note that ethical considerations are an integral part of our methodology, underscoring our commitment to responsible and transparent research practices.

This multifaceted methodology, covering data preparation, preprocessing, and model development, underscores the rigor and comprehensiveness of our study. It allowed us to effectively leverage machine learning techniques, specifically the XGBoost algorithm, to establish a robust framework for the early detection of Parkinson’s disease.

## Results and discussion

Our study has produced impressive results in the development of a classification model designed for early disease detection, particularly for Parkinson’s disease (PD). PD is a progressive neurodegenerative disorder impacting the central nervous system, resulting in a range of motor and non-motor symptoms. Early diagnosis of PD is pivotal for effective disease management. Our model has demonstrated an exceptional level of accuracy, achieving approximately 93.33%, indicating its capability to correctly identify individuals with or without the condition.

Furthermore, our model exhibited a precision of about 91.67%, signifying its proficiency in making precise positive predictions while minimizing the chances of false positives. The model’s recall, an indicator of its sensitivity, was approximately 95.65%, highlighting its effectiveness in capturing the majority of individuals with the condition, consequently reducing false negatives. The F1-Score, which balances precision and recall, also stood at approximately 95.65%, emphasizing the model’s equilibrium between accuracy and inclusivity in its predictions.

The accompanying confusion matrix provided a detailed view of our model’s performance. Among the 30 instances, our model correctly identified 22 individuals with the condition (true positives) and accurately classified six individuals as not having the condition (true negatives). Nevertheless, it did produce one false positive and one false negative prediction. The excellent combination of high precision, recall, and F1-Score signifies the model’s potential in real-world healthcare settings, where early and accurate disease detection is paramount.

Additionally, our model’s robust performance is validated by the high AUC-ROC and AUC-PR scores, approximately 0.9689 and 0.9906, respectively. These scores reinforce the reliability of our model in effectively distinguishing between positive and negative cases and optimizing the precision-recall trade-off.

These results not only underscore our model’s impressive performance but also emphasize its transformative potential in the healthcare domain, especially in the context of early disease detection. With its strong precision and recall, it has the potential to significantly reduce misclassifications and aid healthcare providers in making well-informed decisions. The exceptional AUC-ROC and AUC-PR scores further strengthen the case for our model’s reliability and its ability to differentiate between positive and negative cases.

In conclusion, our model shows significant promise for early disease detection and exemplifies the potential of machine learning in enhancing patient outcomes and healthcare efficiency. It serves as an illustration of the growing intersection of technology and healthcare, inspiring further research, development, and practical implementation in diagnostic and clinical settings. It has the potential to improve the early diagnosis and management of diseases like Parkinson’s, leading to better patient outcomes.

The application of machine learning in the healthcare sector, particularly in the detection and diagnosis of Parkinson’s disease, is a promising and transformative approach. Parkinson’s disease, a prevalent neurodegenerative condition, presents unique diagnostic challenges due to its subtle early symptoms. Machine learning has emerged as a powerful tool to overcome these challenges, utilizing diverse data sources, including voice, image, and motion data, to create effective detection and diagnostic systems.

The reviewed articles from notable researchers (Gomathy et al., Kulkarni et al., Hossain et al., Zhang et al., and Pinto et al., all from 2023) have consistently demonstrated the potential of machine learning in Parkinson’s disease detection. These studies reported high accuracy levels, with some achieving up to 97% accuracy in detecting PD using voice data. A meta-analysis conducted by Zhang et al. confirmed the consistent high accuracy of machine learning systems, emphasizing their potential to detect PD with 90% accuracy or higher.

However, it is essential to recognize that while these results are promising, machine learning systems are still evolving and should not replace professional medical diagnosis. Individuals with suspected Parkinson’s disease symptoms should always seek evaluation and diagnosis from healthcare professionals. Additionally, the effectiveness of machine learning systems may vary depending on data types, machine learning methods, and the quality and size of the datasets used for training and evaluation.

In summary, the reviewed studies provide encouraging evidence of machine learning’s capacity to revolutionize Parkinson’s disease diagnosis and management. Further research and validation are needed to develop robust and generalizable machine learning systems for diverse patient populations, but the potential benefits for both patients and healthcare professionals are substantial.

## Conclusion

In this study, we have showcased the potential of machine learning, particularly the XGBoost algorithm, in the detection of Parkinson’s disease. Our rigorous methodology, involving data curation, preprocessing, and model development, has led to a highly accurate and promising framework for early disease detection.

Our model achieved an impressive accuracy of approximately 93.33%, demonstrating its ability to correctly identify individuals with or without Parkinson’s disease. With a precision of about 91.67%, our model excelled in making precise positive predictions, minimizing the likelihood of false positives. The model’s recall, at approximately 95.65%, highlighted its effectiveness in capturing the majority of individuals with the condition, reducing false negatives and ensuring sensitivity. The balanced F1-Score, also around 95.65%, emphasizes the model’s equilibrium between accuracy and inclusivity in its predictions.

A detailed examination of our model’s performance through the confusion matrix revealed its capability to correctly identify 22 individuals with the condition (true positives) and accurately classify six individuals as not having the disease (true negatives). While there were isolated instances of one false positive and one false negative prediction, the overall performance, characterized by high precision, recall, and F1-Score, underscores the model’s potential in real-world healthcare settings.

The model’s robust performance is further substantiated by the impressive AUC-ROC score of approximately 0.9689 and AUC-PR score of about 0.9906. These high scores reinforce the reliability of our model in effectively distinguishing between positive and negative cases, optimizing the precision-recall trade-off and minimizing the potential for misclassification.

These numeric results, combined with the well-established capability of machine learning in the context of Parkinson’s disease detection, emphasize the transformative potential of our model in the healthcare domain. The strong precision and recall achieved by our model have the potential to significantly reduce misclassifications and support healthcare providers in making well-informed decisions. The high AUC-ROC and AUC-PR scores further strengthen the case for our model’s reliability and its ability to differentiate between positive and negative cases with precision.

In conclusion, our study demonstrates the significant promise of machine learning, particularly the XGBoost algorithm, for early disease detection. The potential benefits for both patients and healthcare professionals are substantial. As we move forward, it is imperative to continue refining our model and consider validation on a larger and more diverse dataset. Parkinson’s disease detection remains a complex task, and further research and clinical validation are essential for real-world applications. However, our model serves as a beacon of hope in enhancing patient outcomes and healthcare efficiency, especially in the context of diseases like Parkinson’s.

## Data Availability

All data produced in the present work are contained in the manuscript

